# Lower probability of viral suppression in people living with HIV who are parents in Lima, Peru

**DOI:** 10.1101/2024.01.04.24300848

**Authors:** Valeria Navarro-Galarza, Elsa González-Lagos, Jorge Robledo, Ana Graña, Eduardo Gotuzzo

## Abstract

**Introduction:** Parenting can be a detriment for selfcare among people living with HIV (PLWH) out of concern for their children’s futures and responsibilities that may limit attendance to health services. We explored the association between having children <20 years-old and viral suppression in PLWH.

**Methods:** Retrospective cohort study from secondary data of PLWH enrolled at the largest HIV program in Lima between 2012-2018. We established parenthood by participant self-report children <20 years-old at enrollment, with additional data gathered for PLWH who reported a child born during the first year after enrollment. The main outcome was viral suppression (<400 copies/mL) by the end of follow-up. We conducted Cox regression analyses for repeated events, censoring at time of death or lost to follow-up. We built the final model by backward stepwise regression including potentially important variables and those with p-value ≤ 0.20 in bivariate analyses, presenting hazard ratios (HR), adjusted HR (aHR), and 95% confidence intervals (CIs).

**Results:** In 3170 PLWH, median age at enrollment was 31.6 years (range 17.9-76.1), 79.8% were men and 27.2% reported having children <20 years-old (median=2). At end of follow-up (8766.6 person-years), 534 (62.0%) were virally suppressed. In our final multivariate model, having children <20 years-old (aHR 3.53; [95% CI] 1.88 - 6.62) and the birth of a child during the first year after enrollment in the program (HR 1.81; [95% CI] 1.30 – 2.50) were independently associated with lack of viral suppression by end of follow-up. Based on health status of the PLWH, we estimated 70% and 69% of children to be at risk of maternal and paternal orphanhood, respectively.

**Conclusion:** In our setting, being a parent was associated with lower probability of viral suppression, creating a risk of orphanhood for children of PLWH. Family support services may facilitate HIV care and help PLWH maximize viral suppression.

## Introduction

Parenting is a transforming life experience that demands time-consuming responsibilities to assist children [1]. Parents affected by chronic diseases may struggle to balance their own health care needs with parenting responsibilities. Despite HIV predominantly affecting people of reproductive age, there have been few studies regarding parenthood effects on parents’ HIV health outcomes [2]. Studies have pointed to responsibilities of daily living, including parenting, as potential barriers for retention in care of people living with HIV (PLWH) [3–5]. Others suggest negative associations between parenting of PLWH and positive children’s outcomes. [6–8].

Parenting-associated effects may depend on a complex interplay of individual and societal factors. Child and family factors include age, health status, and number of children. Factors for the parent living with HIV include sex, the availability and quality of partner and social support, economical means, gender disparities, and the provision of health services.

Women typically bear more responsibilities for children in their social, biological, and occupational roles, resulting in a greater family and childcare responsibilities. In Peru, 40.3% of women between 15 to 24 years reported interruption of studies due to familial or economic reasons, including pregnancy, and care of young children [9]. Lower adherence to antiretroviral therapy (ART) in women compared to men has been described [10]. A Lima study showed 47% of women ART dropouts [11]. In a multinational study in Latin America, women were overrepresented among PLWH who were deceased or lost to follow-up [12]. A 2014 study reported 40% of lost to follow-up (LTFU) of women living with HIV in their first year after newborn delivery [13].

In Peru, health service providers may not appreciate that PLWH may have substantial childcare duties that inhibit their own HIV-related care. In the absence of optimal ART to maintain viral suppression, premature parental death may result in orphanhood for children of PLWH. Orphanhood can inhibit child development [14], further aggravating the societal impact of HIV.

We studied the association between parenthood, defined by self-report of having children <20 years of age, with viral suppression by the end of follow-up in PLWH in a Lima, Peru, HIV clinic. We also studied orphanhood and the risk of orphanhood for children of PLWH.

## Methods

### Study setting

In Peru, 24% of PLWH were men who have sex with men (MSM) and has not yet met UNAIDS continuity of care goals (86-82-61) by 2022 [15]. In women, an HIV infection is commonly identified in their reproductive age during pregnancy due to university HIV testing in antenatal care or in family planning consultation [16]. In men and transwomen, campaigns, outreach, and screening programs targeted MSM are efforts tailored to national guidelines [17].

Our study analyzed data from the largest HIV Program in Lima. Based at a public hospital in the north of Lima, it enrolls people aged 18 years and older with a 3^rd^ or 4^th^ generation ELISA HIV reactive test result followed by a confirmatory test and viral load.

During the enrollment visit, the nursing staff provides individualized counseling and completes a standardized individual form with general and clinical information. A multidisciplinary team comprehensively evaluates circumstances of PLWH enrolled in the program before the start of ART, which is free at public hospitals. Eligibility guidelines during the 2012-2017 study period including all persons who were pregnant, had tuberculosis, or had CD4+ cell counts <350 cells/mL.

Clinic nurses perform clinical evaluations every 6-9 months for viral load and CD4+ cell counts tests. Medications are obtained either by the PLWH or a designated person at additional visits scheduled by the nursing staff. Patient data is provided to the infectious disease specialists during outpatient visits. Demographics data is entered into the HIV Program database with a unique code, updated after each visit. Separately, outpatient visit data is recorded in the outpatient database with the same code.

During the enrollment visit, we invited adult PLWH to participate in an HIV Cohort Study at the study center. For those who consented, the entry record of the Cohort Study verified the sociodemographic and clinical information and collected data on partners and children (number and ages). Cohort data were enhanced with study-specific information described below.

### Study design

We based our retrospective cohort study on the integration of secondary data of PLWH enrolled at the HIV program of our study center in Lima. The main predictive factor of interest was whether a PLWH reporting having one or more children <20 years of age at the time of enrollment in the HIV Program. The main outcome was viral suppression at the end of follow up.

### Study population

PLWH were eligible for study inclusion if they were enrolled into the HIV Program between January 1, 2012, and December 31, 2017, and were themselves at least 18 years old with a confirmed HIV positive serostatus. The study follow-up was closed on December 30, 2018.

### Data sources

We merged four datasets: the HIV Program individual forms, Cohort Study, nursery records and the outpatient service records. From the HIV Program forms, we identified the eligible study population and extracted general and clinical information, including age, sex, date of HIV diagnosis, World Health Organization (WHO) AIDS stage, CD4+ cell count, viral load, ART regimen, pregnancy, ART pick-up dates, and vital status by the end of follow-up. From the outpatient service database, we obtained the dates of outpatient visits, which served to document retention in care. From the Cohort Study, we obtained data about participants’ partners, partners’ HIV status, parental status, and the number and ages of living children. From nursing queries and HIV clinic records, we obtained the information of the newborn children. Data was retrospectively collected during the second and third quarters of year 2019.

### Study definitions

Under WHO classification, we considered infants/children to be 0 to 9 years old and older children/adolescents from 10 to 19 years old. [18]. The main outcome, viral suppression, was defined as viral load detection <400 copies/mL in a measure at least 180 days after enrollment; the cut-off was consistent with the laboratory detection limits in the study period [19]. Retention in care was determined by a person’s attendance at two medical visits in a period of more than ninety days, as per the U.S. Health Resources and Services Administration definition.

We considered PLWH to be LTFU if they informed staff that they left the program or if they fulfilled program abandonment criteria, defined by the Peruvian Ministry of Health as PLWH whose ART is not picked-up by the person themselves or by the designated person for more than 30 consecutive days (ART was given out monthly in the 2012-2017 study period) since the scheduled pick-up date [20, 21].

For the children of PLWH, the vital status of the parent with HIV by the end of follow-up defined HIV orphanhood. We considered death for single-parent HIV orphanhood, and the following variables as risk of orphanhood due to HIV: PLWH who abandoned ART, lack of viral suppression, or lack of retention in care.

### Data management and analysis

We merged the four datasets based on the HIV Program’s unique patient code, with verification of patient initials and month/year of birth. Personal identifiers were not stored in the final dataset. In the data cleaning process, we checked for duplicate records (i.e., the same individual enrolled twice at different times) and inconsistent information, especially for the number and ages of children; inconsistencies were found in <0.5% of records and resolved by reviewing the source documents. Persons without reliable data on parenthood were excluded.

We also assessed the proportion with childbirth during the first year of follow-up and the proportions with viral suppression or who were dead at the end of follow-up. Since it was not possible to identify children who could have both parents in the study, the percentages of single-parent (at least) orphanhood and children of living PLWH at risk of orphanhood at the end of the follow-up were presented according to the sex of PLWH.

For the statistical description of categorical variables, we used frequencies and percentages. For continuous variables, we used measures of central tendency and dispersion according to the observed distribution of values, i.e., for normally distributed data, we used means with standard deviation and for skewed data, we used medians and interquartile ranges. For inferential statistics, we used bivariate analyses and multivariate models. In the main bivariate analyses, we compared the proportion of viral suppression by the end of follow-up per groups defined by the presence of children <20 years of age. For the analysis of the independent and adjusted effects of our exposure on viral suppression, we used a Cox proportional hazards model adapted for repeated events with death and LTFU as censored events. In this model, we included all potentially significant variables, either due to results in bivariate analyses (p <0.2), or clinical criteria; we also included interactions between PLWH older age (≥50 years) and having partners with children <20 years of age due to their social burden relevance. The strength of the association was estimated with Hazard Ratios (HR) and 95% confidence intervals (95% CI). P-values are considered statistically significant when <0.05, two-tailed.

### Ethics statement

The Ethics Committee of Universidad Peruana Cayetano Heredia and of the study center reviewed and approved the study protocol. We used HIV program information as adapted to the personal data protection regulations of Peru [22], deploying good data management practices to preserve confidentiality [23].

## Results

Between 2012 and 2017, the clinical records of 3170 of 4051 PLWH (78.3%) enrolled in the HIV program of the study center indicated that subjects met our inclusion criteria and were included in the study (**Figure 1**).

**Fig 1.**
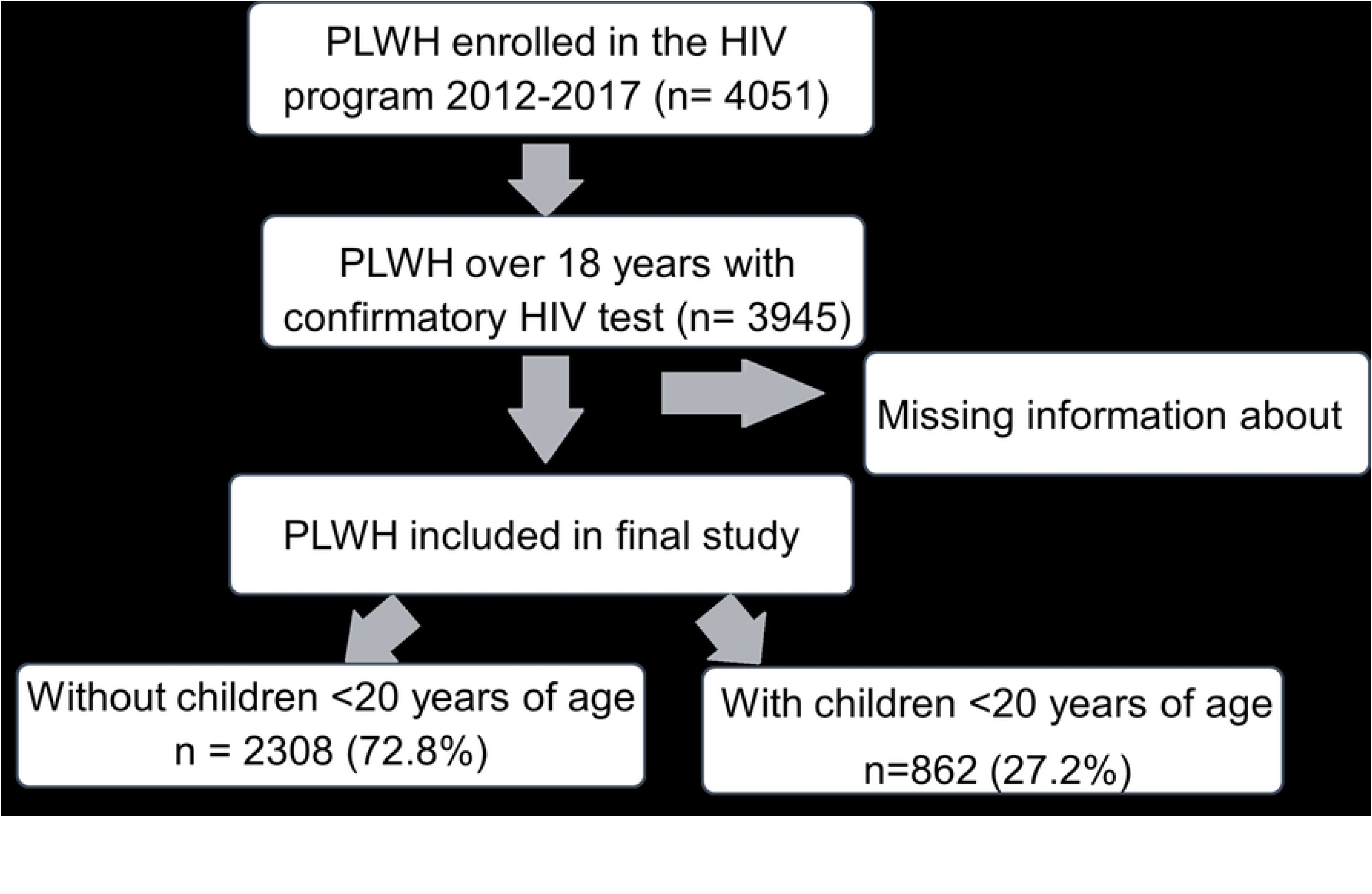
Flow chart of the clinic population of persons living with HIV (PLWH) included in the parentage-viral suppression study in Peru.

The 3170 PLWH had a median age of 31.6 years (interquartile range [IQR]: 25.4-40.3) and men accounted for 79.8%, similar to data from the 688 PLWH in the HIV Program who were excluded due to missing parental data (median age 31.7 years; 78.2% men). Data on residence indicated that 1399 (44.1%) came from districts outside the clinic catchment area. Almost half (45.2%) of men self-reported as MSM and most women as heterosexual (90.5%). Stable sexual partners were reported by 34.7% of men and 70.1% of women.

At enrollment, 27.2% (862/3170) of PLWH reported being parents of children <20 years of age (median=2 [IQR: 1-3]; **Table 1**). Of the 978 men with partners, 32.5% (318) had children <20 years of age, and of the 427 women with partners 67.0% (286) had children <20 years of age. For PLWH without children at any age stage (n=2135), 15.8% (337/2135) reported having a stable partner with HIV infection. Men were less likely to report having a partner with HIV than women (17.0% vs. 32.7%). At their first HIV Program clinic visit, 51.1% (1620/3170) of the participants were <1 month of their initial HIV diagnosis. The median CD4 count at admission was 248 (IQR: 97-428), men’s was 243 (IQR: 94-415.5) and women’s was 269 (IQR: 113-461). 47.4% (1504/3170) were in WHO stage 2, 46.3% (1173/2530) of men and 51.7% (331/640) of women were in the same stage.

**Table 1.**
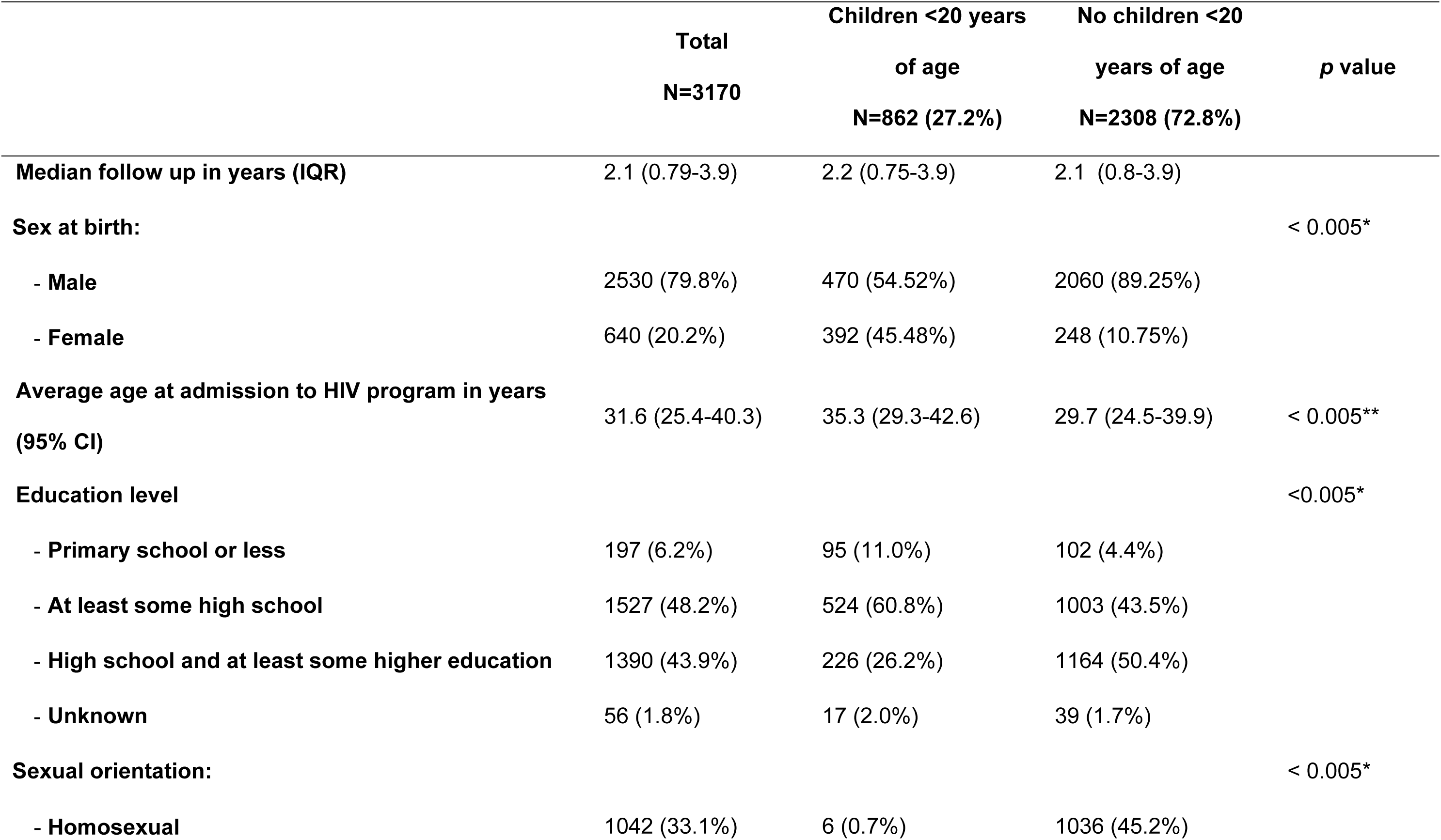
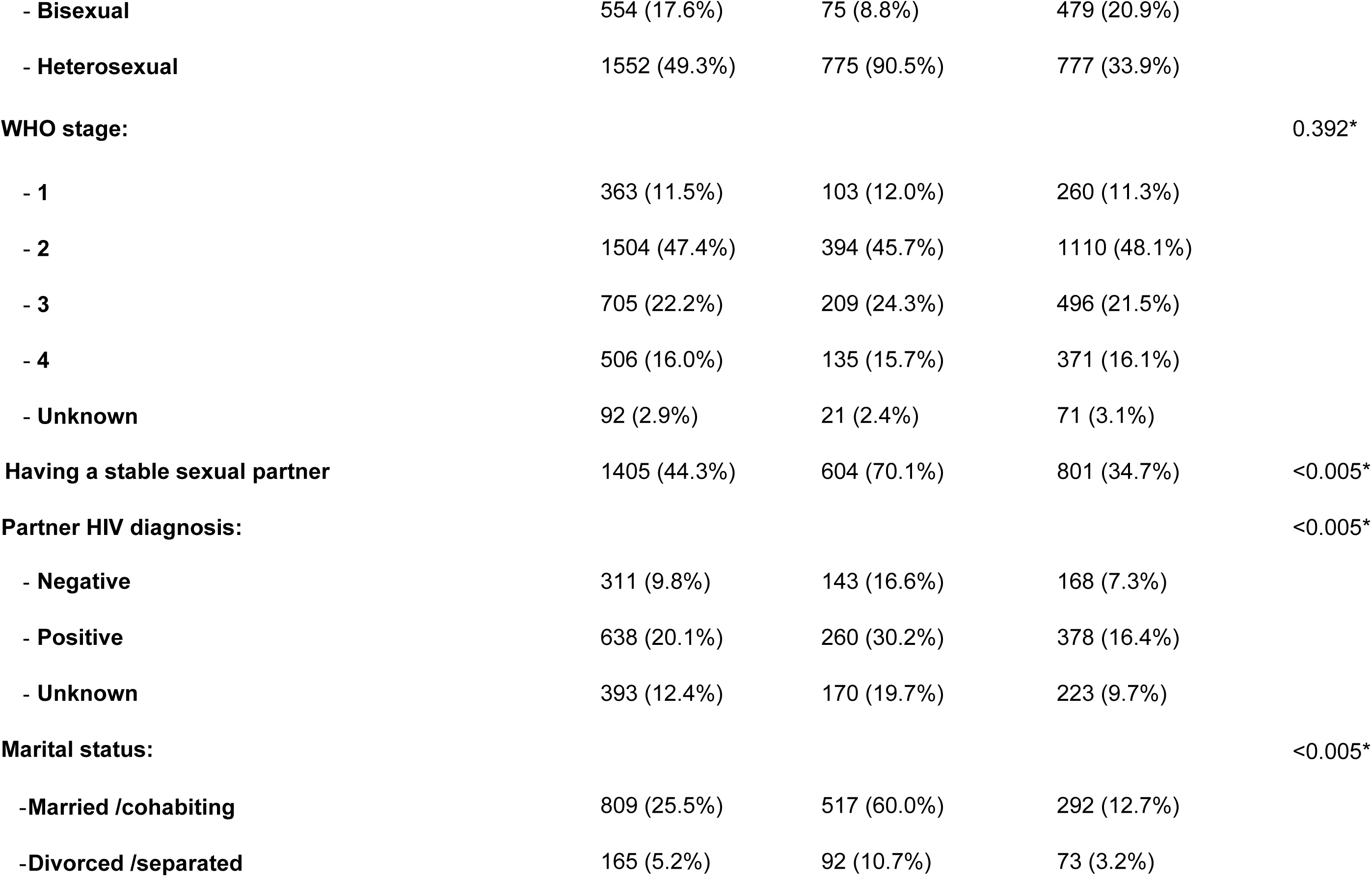
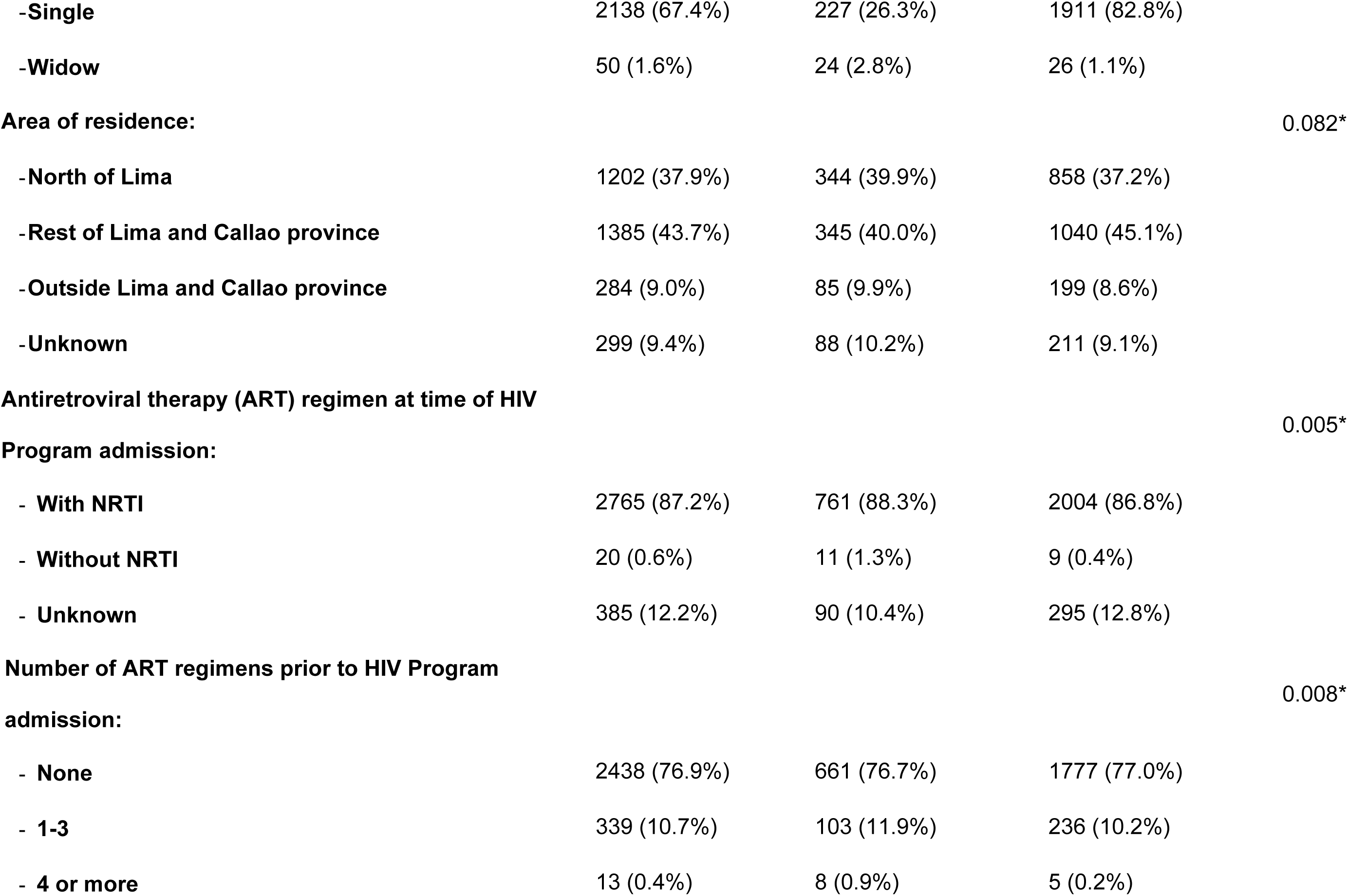
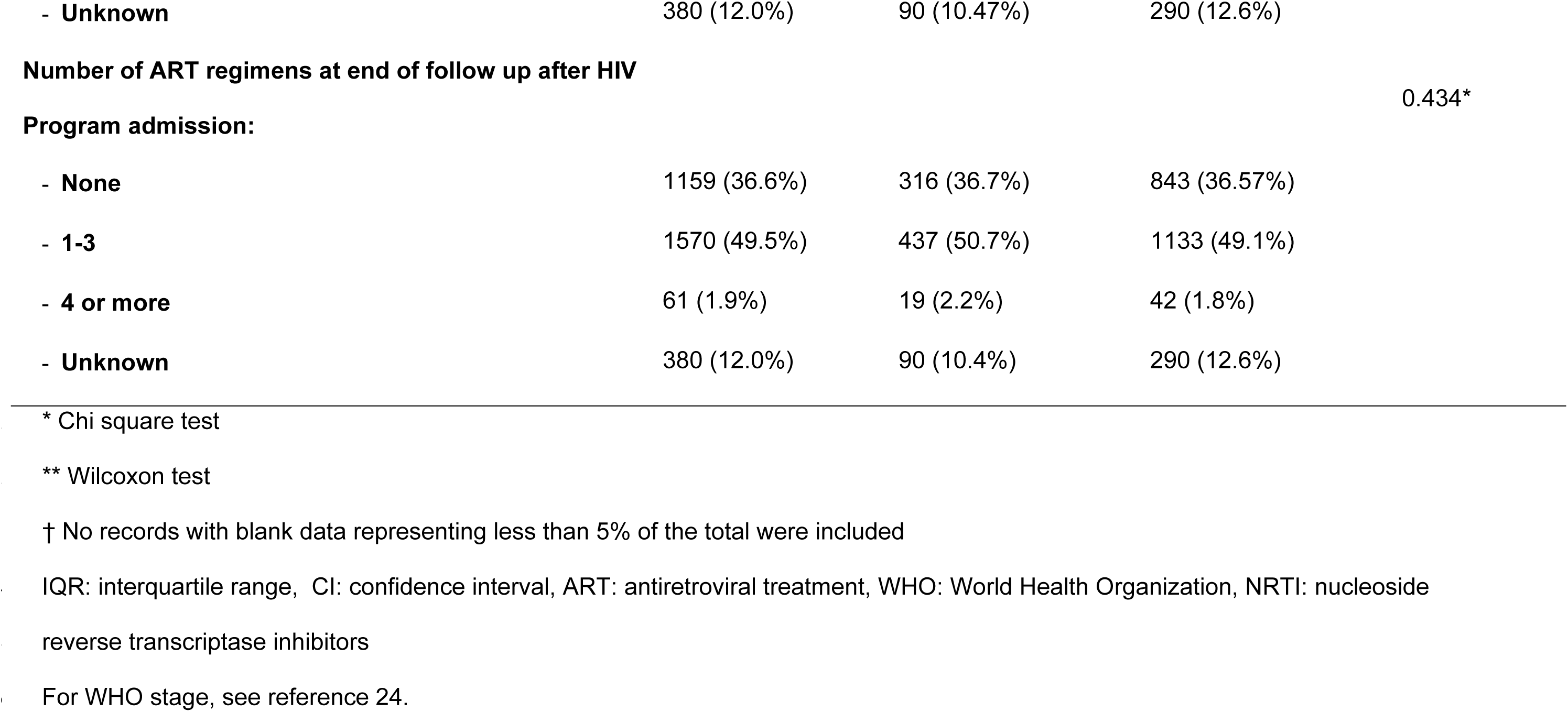
Baseline characteristics of the 3170 persons living with HIV by parental status.

At time of HIV Program enrollment, 88.1% (2792/3170) participants were started on ART, 99.0% (2765/2792) with regimens containing nucleotide reverse transcriptase inhibitors. 56.2% (1570/2792) changed ART regimens, sometimes more than once. In 3.4% of PLWH (109/3170) the birth of a child was identified during the first year of follow-up, 89% (97/109) in women. 80.1% (2540/3170) of people had a viral load measurement made at least 180 days after the first visit and 65.5% (2075/3170) achieved viral suppression at the end of the follow-up. Of the 862 PLWH with children <20 years of age, 61.9% (534/862) achieved viral suppression at the end of follow-up. 6.9% (218/3170) of PLWH died during follow-up; of whom 30.3% (66/218) had children <20 years of age.

The final survival analysis includes 3129 PLWH applicable data records for non-viral suppression at end of follow-up (**table 2**).

**Table 2.**
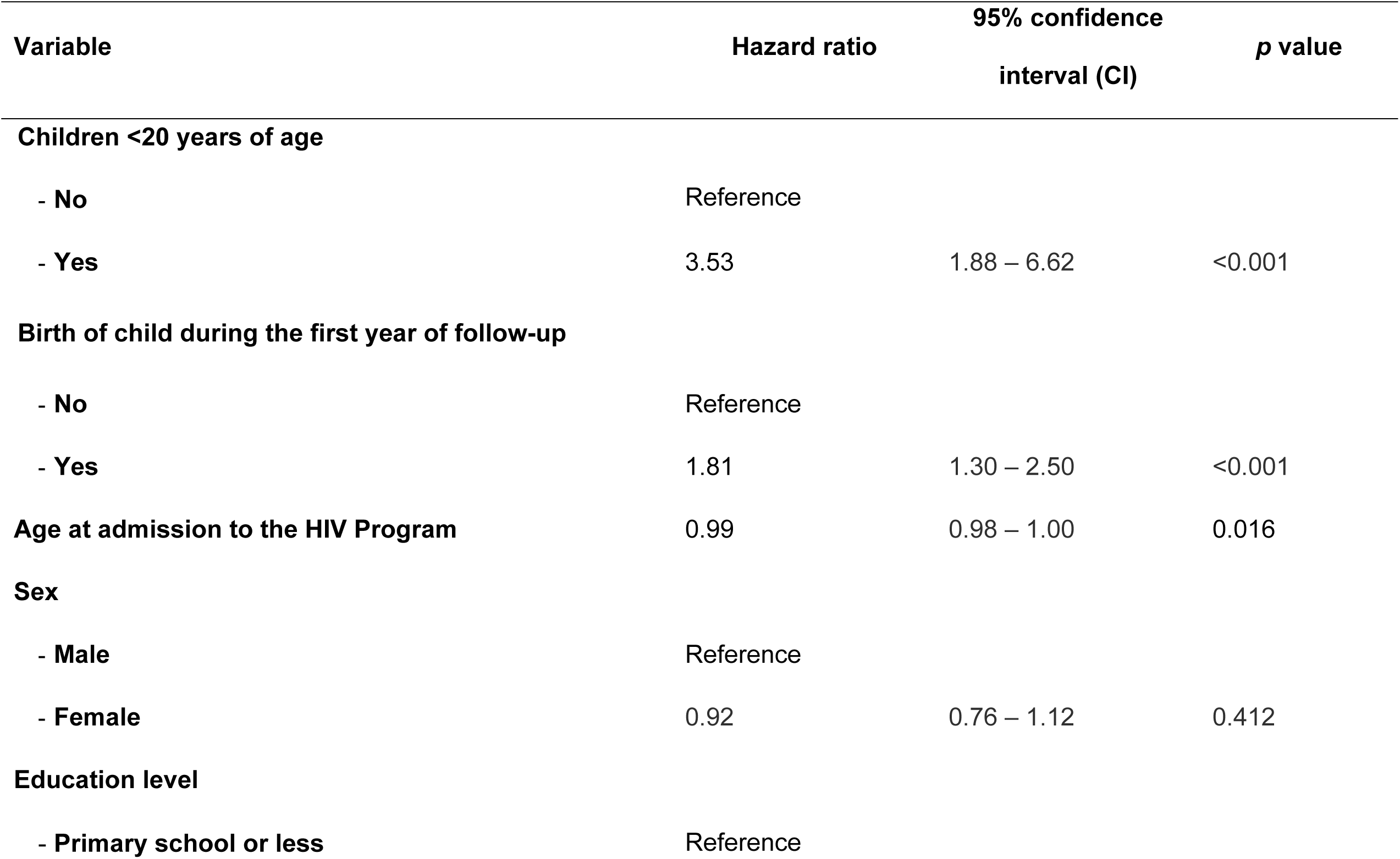
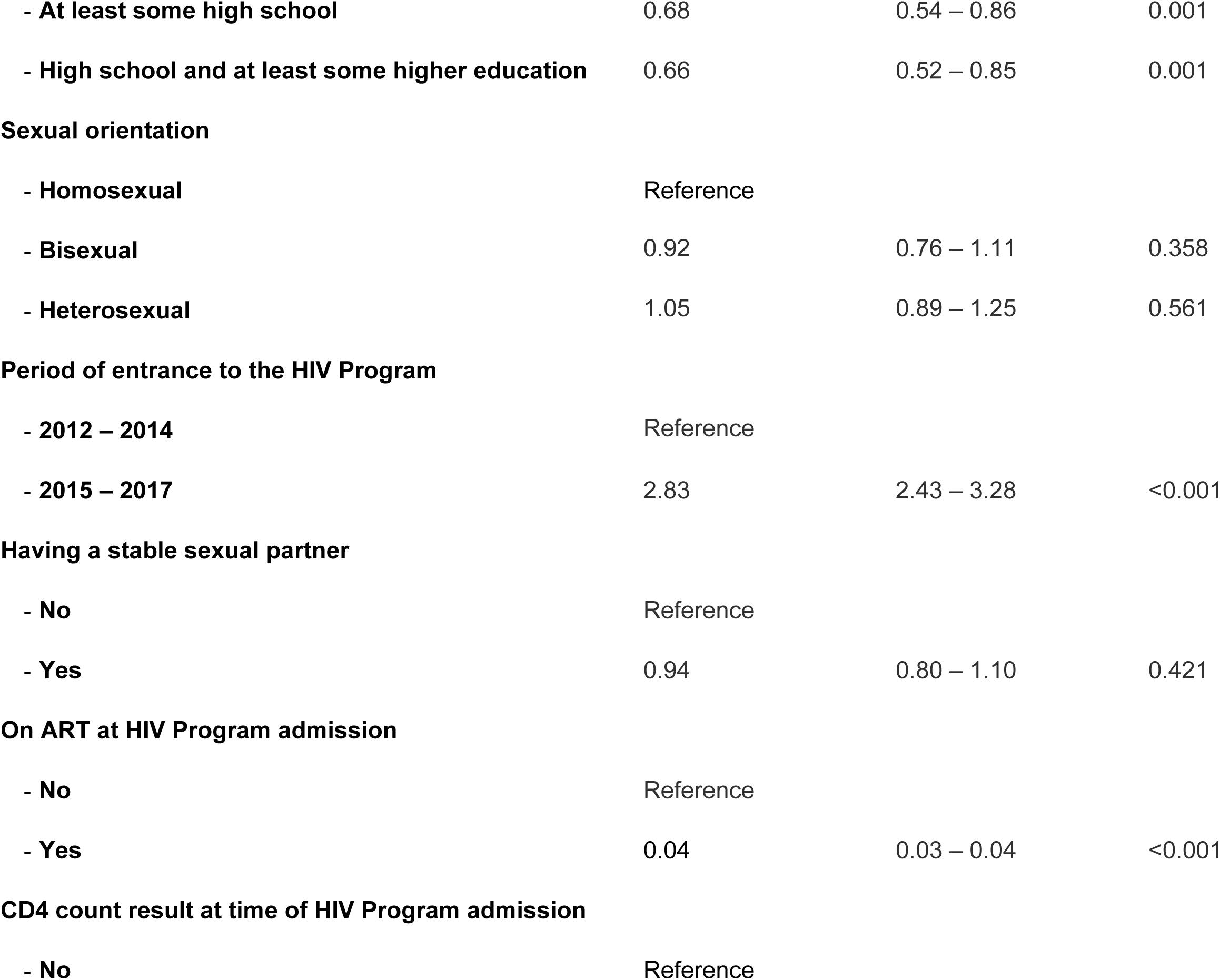
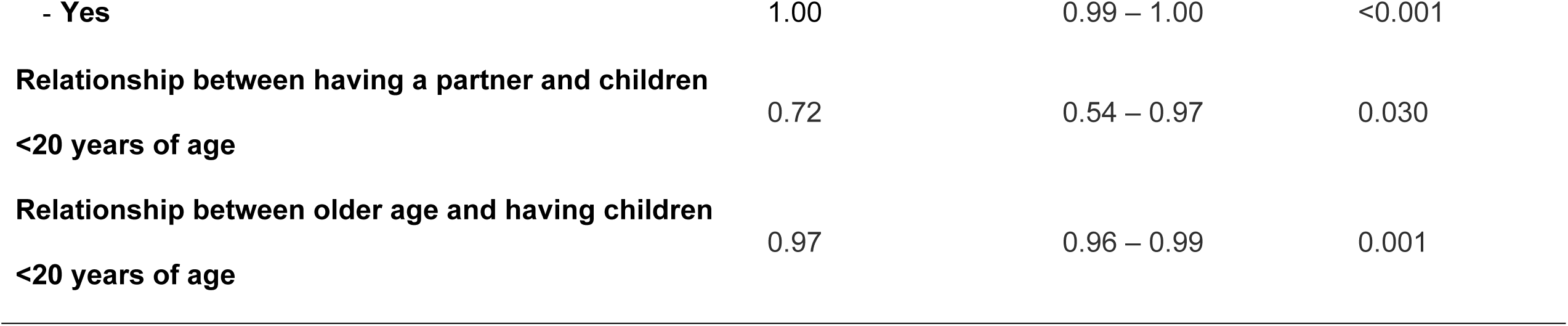
Survival analysis for the outcome viral non-suppression at the end of follow-up of the study for 3129 persons living with HIV.

**Figure 2** shows the overall less likelihood of achieving viral suppression at the end of the follow-up of PLWH with children <20 years of age in comparison to those without children <20 years of age. **Figures 3 and 4** show that women with children <20 years of age are less likely to achieve the outcome of interest in comparison to women without children <20 years of age; in contrast, men in both groups follow almost the same pattern of viral suppression over time. In **figure 5**, the classification by having children above or under 20 years-old evidence that the group with younger children were less likely to achieve viral suppression in comparison to the rest of the groups; the one with older children only had better performance at the end of the follow-up.

**Fig 2.**
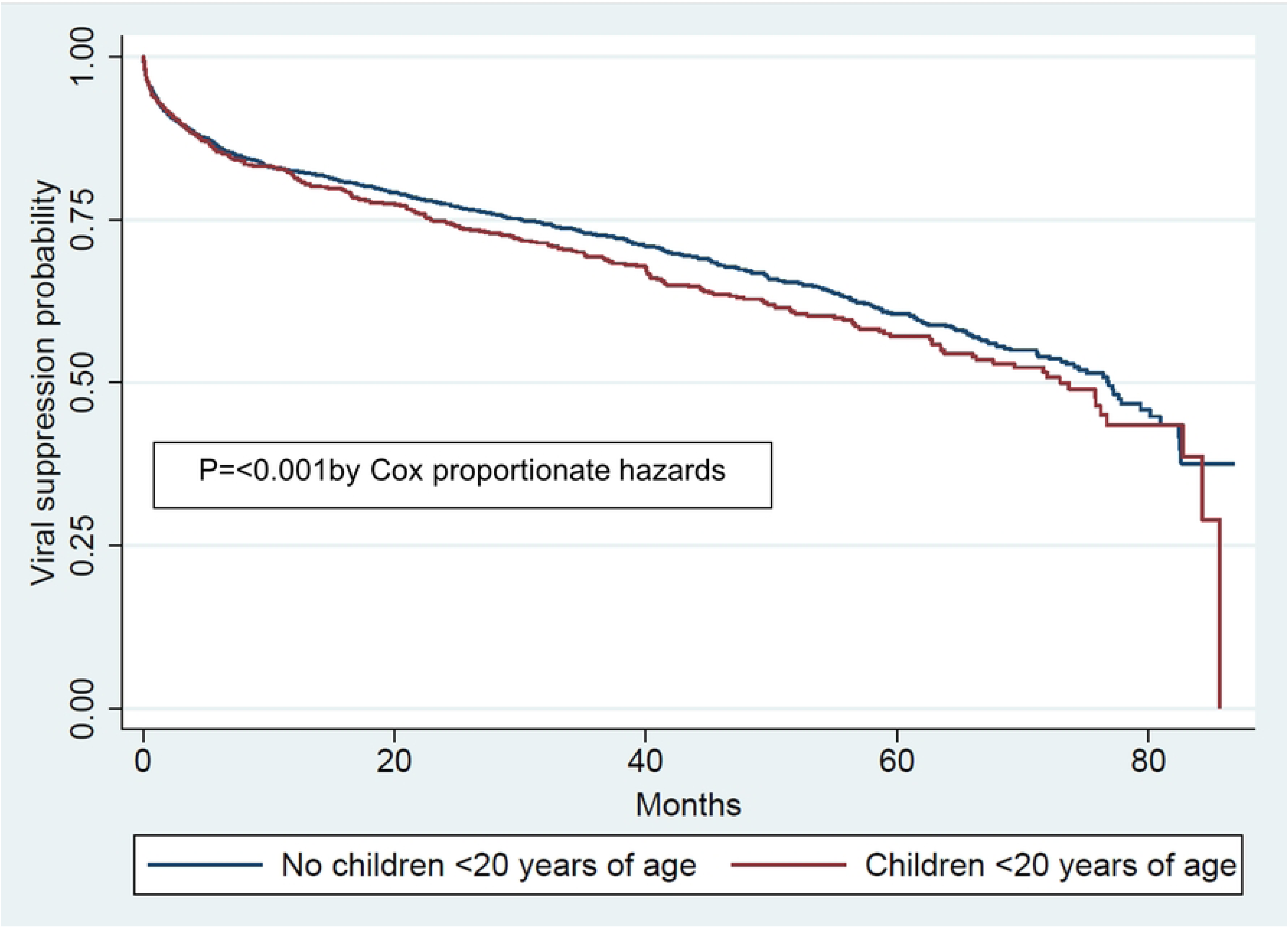
Kaplan-Meier curve: viral suppression outcome at the end of follow-up of the study by having children <20 years of age for 3129 persons living with HIV.

**Fig 3.**
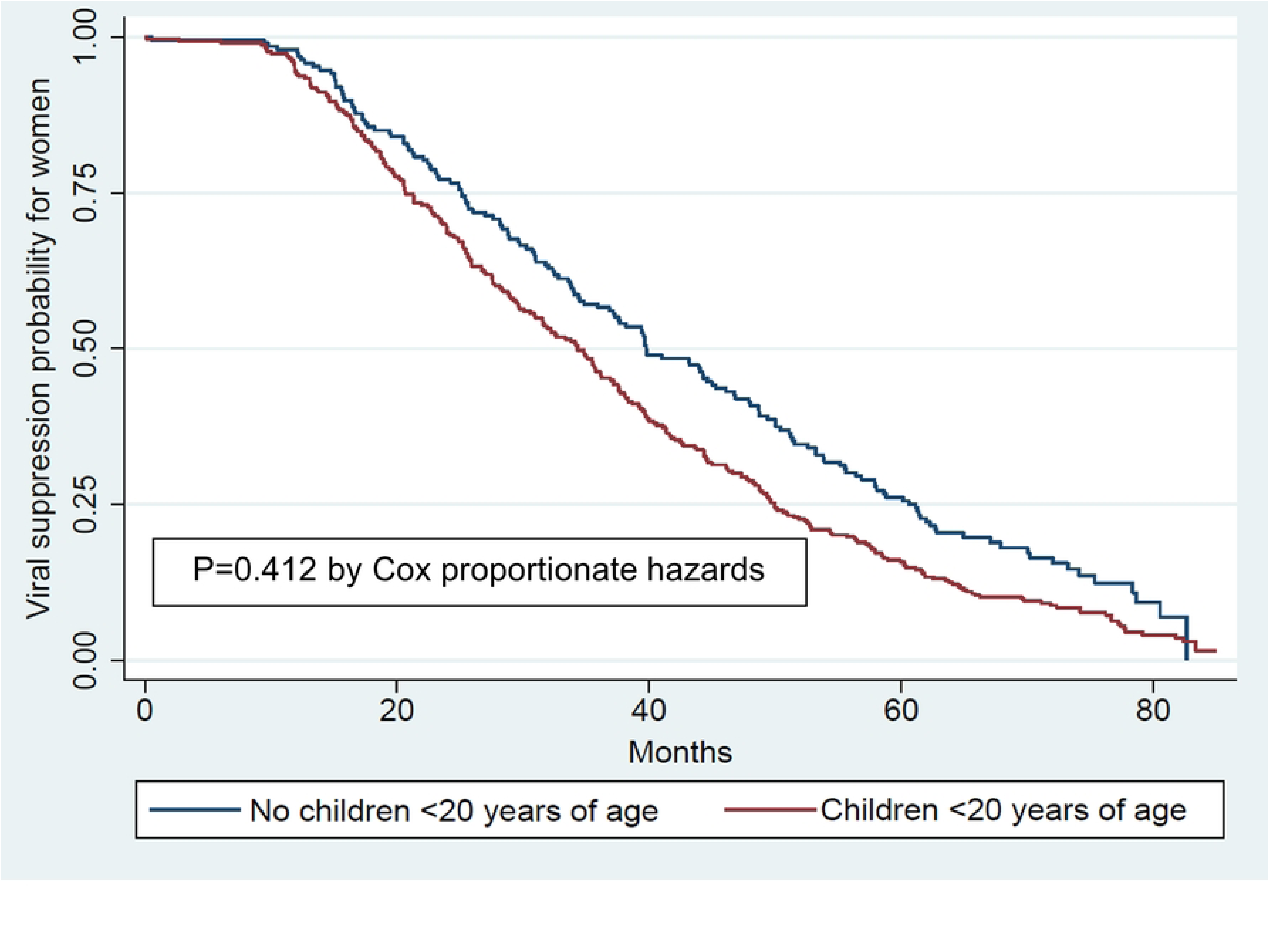
Kaplan-Meier curve: viral suppression outcome at the end of follow-up of the study by sex, classified by having children <20 years of age for 3129 persons living with HIV.

**Fig 4.**
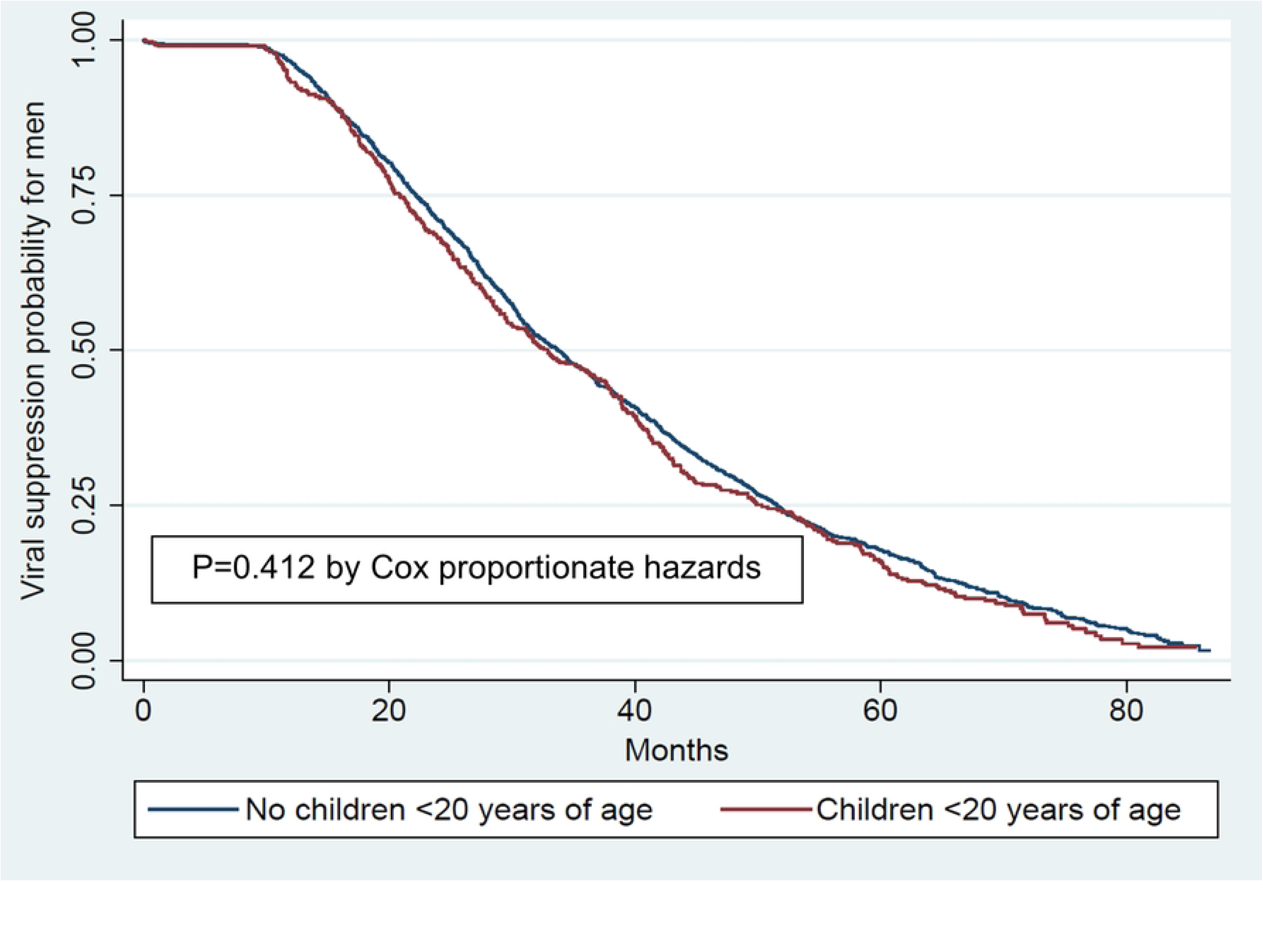
Kaplan-Meier curve: viral suppression outcome at the end of follow-up of the study by sex, classified by having children <20 years of age for 3129 persons living with HIV.

**Fig 5.**
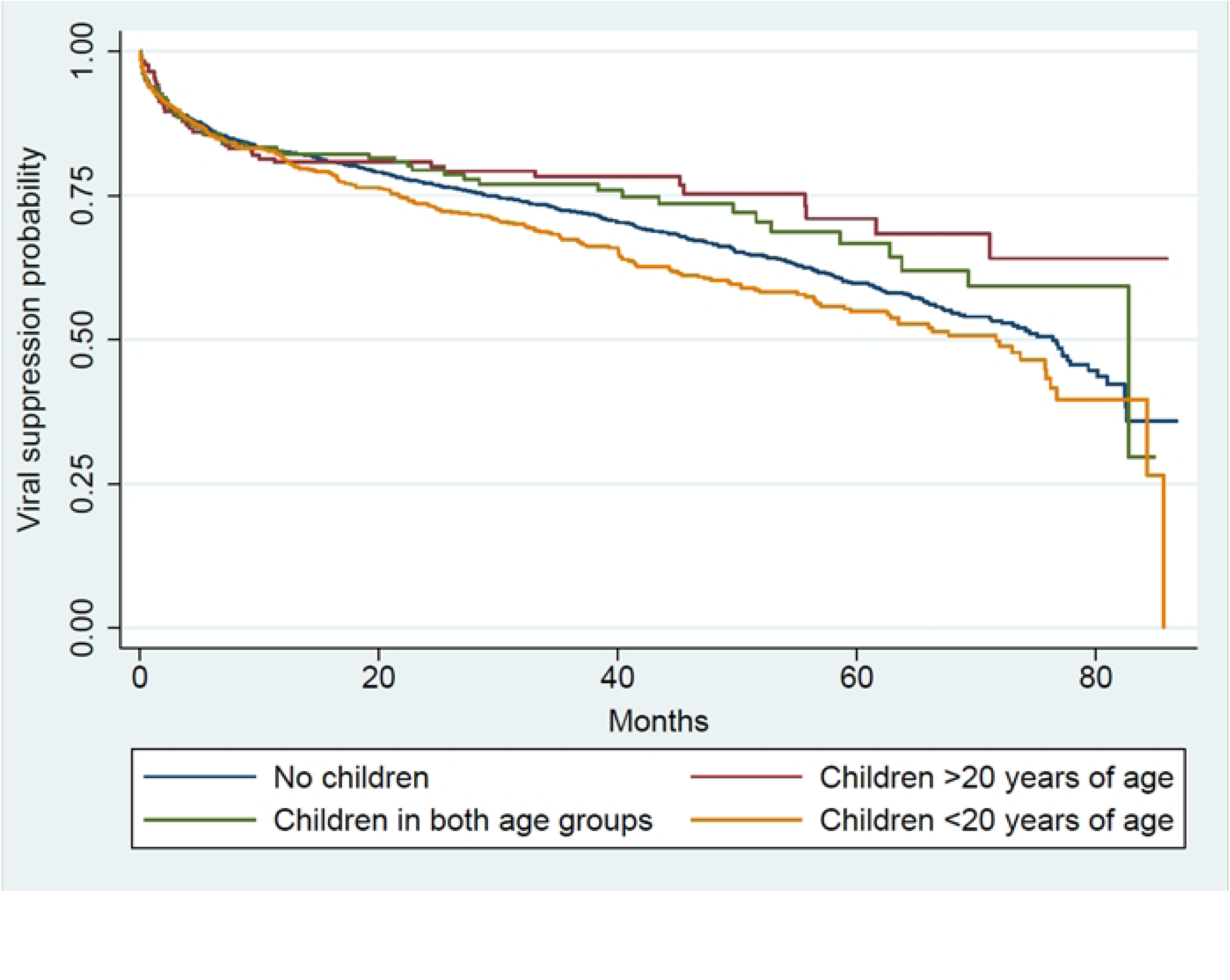
Kaplan-Meier curve: viral suppression outcome at the end of follow-up of the study by children age groups for 3129 persons living with HIV.

At the end of the follow-up (median: 2.1 years), we estimated that 358 PLWH were mothers of 578 children <20 years of age and 428 PLWH were fathers of 674 children <20 years of age; because of the anonymity of the information and the possibility of both parents’ inclusion in our population, we could not estimate the total number of children. By the end of follow up, orphanhood affected 4.3% (25/578) of children <20 years of age of mothers with HIV, and 11% (74/674) of children <20 years of age of fathers with HIV. 69.6% (402/578) of children of mothers with HIV and 69.3% (467/674) of children of fathers with HIV were at risk of orphanhood. At the end of the follow-up, 31.1% (270/869) of children <20 years of age at risk of orphanhood and 37.4% (37/99) of orphan children <20 years of age were 5 years old or less. Women with HIV placed their children at risk of orphanhood due to non-retention in care (140, 50.5%), non-viral suppression (118, 42.6%) or ART abandonment (19, 6.9%). Men with HIV placed their children at risk of orphanhood due to non-retention in care (178, 56.7%), non-viral suppression (124, 39.5%) or ART abandonment (12, 3.8%).

## Discussion

In our clinic-based study in Lima, Peru, PLWH with children <20 years of age were less likely to achieve viral suppression by the end of 2+ years of follow-up. This was also the case for PLWH who reported the birth of a child during the first year after enrollment. Our population had demographic characteristics similar to PLWH whose parental status was unknown, suggesting the generalizability of our findings.

Our hypothesis was that PLWH parenthood may be a stress that paradoxically increases parental health vulnerability. We found evidence that, in fact, PLWH who are parents had less probability of viral suppression. In front of the infants’ needs, those of the parents might be overlooked, postponed or, even, not addressed [25]. This situation poses additional health implications for people living with chronic diseases, including HIV [26–28], particularly if the social support is limited or access to health services is burdensome. During the postpartum period, greater anxiety, fatigue, depression and decreased self-care are frequent for women [29, 30]. Men are also prone to depression, especially if the partner is depressed [31], and to anxiety related to their own role as a father and to the economic factors that influence their family stability [32–34].

The proportion of PLWH with children <20 years of age by time of enrollment in our HIV program was substantially higher in women compared to men. In Peru, the laws still limit LGBT parenting, which may explain our results. Women are still the main caregivers of children across cultures; however, our results suggest that parenthood of children <20 years of age would also affect HIV care in men, although to a lesser extent. A parallel study at our study center found that women showed less adherence and retention in care in puerperium rather than during pregnancy [35]. Figure 3 shows less likelihood of viral suppression in women having children <20 years of age in comparison to the ones without them, whereas results remain almost the same for both men groups. Parents of young children tend to be young, which has been related to HIV care interruption and lost to follow up [36]. These aspects emphasize that parental health vulnerability of PLWH should be understood and not dismissed, with a special focus on women. In our study, pregnancy was the most frequent reason for HIV diagnosis in women. In many cases, childbirth occurs during the stage of initial coping thus probably prior to disclosure to the trusted extended family, reducing the offer and request of support due to stigma [37–40]; however, we did not collect disclosure data. This aspect might explain the association between non-viral suppression and childbirth during the first year of follow-up described. It could also explain the results in Figure 5 that shows a difference between the children ages groups: PLWH whose diagnosis was done and disclosed years ago might be more likely to have a strongest support network; 75% of our population with children >18 years old were enrolled in the HIV program for more than three years.

Programs worldwide have overlooked HIV-related interventions targeted to the PLWH’s children and adolescents who might need to receive education on HIV-related topics and mental health services [41]. WHO have recommended differentiated service delivery responsive to the preferences and needs of PLWH, including those of parenting [42]. This calls for HIV programs whose services display flexibility towards parental and family needs. At our setting, the HIV National Guidelines demanded multidisciplinary visits before ART start for most of the study period [20, 21]; however, the administrative systems only authorized one visit per day. Complying with multiple visits may particularly challenge people with children under care and limited social support.

Because threats to parental health put their children at risk, we also analyzed orphanhood and risk of orphanhood for the children <20 years of age in our study population. We found that the percentage of orphans <20 years of at least one HIV parent exceeds the 6.6% national average [9]. Orphans of PLWH tend to have greater depression, anxiety, adaptation problems, and learning difficulties compared to orphans due to other causes, which may reduce their future opportunities [43–45]. The surrogate caregivers’ role is fundamental in mitigating negative effects [46].

By the end of follow-up, almost 60% of the children <20 years of age of our PLWH study population were at risk of orphanhood, mainly because of no retention in care of their parents. Due to their parents’ suffering and assuming the duty to care for them, these children may encounter more consequences than actual orphans [44]. A well-established program should create a better household environment that benefits both the individual and their children. The country has in place policies for children in poverty but not for children of sick people at risk of death. The realization of children’s entitlements primarily depends on families, but states have the duty to ensure such rights through adequate social protection [47]. Thus, our results suggest the need of specific health and social protection programs for PLWH and their children.

The study limitations include selection bias due to data from a Cohort Study. However, we did not find major demographic differences in comparison with the non-participant HIV program population. We did not directly control the databases collection procedures; but they are systematically applied by trained nursing personnel, and we conducted comprehensive data management to limit inconsistencies. Valuable covariables such as disclosure, adherence, or mental health were not available to use. Our database does not differentiate PLWH who are considered lost to follow up per the retention in care definition but continue to pick up their ART. We did not determine the total number of children in orphanhood and risk of orphanhood nor the HIV status of the children. The study center corresponds to a reference HIV clinic, which may limit the extrapolation of conclusions. As the main strengths of our study, we gathered data for a large study population with an average follow-up of 2+ years to evaluate the potential impact of parenthood in the care of PLWH and the situation of orphanhood and risk of orphanhood of their children <20 years of age.

Future research in Peru should address the effects of orphaned and at-risk children of parents with AIDS, in addition to evaluating other causes of viral non-suppression related to parenthood. We recommend that the next generation of HIV programs should implement family-centered services, a vision which may further enrich their societal impact.

## Conclusions

The presence of children <20 years of age can have a negative effect on viral suppression in PLWH; likewise, children with PLWH are at high risk of orphanhood. The confluence of increased parental health vulnerability in a context of limited social support and bureaucratic health services may work against continuity of HIV care and threat parental health, which ultimately creates risks also for the children. HIV programs should adapt their services considering individual and family needs of PLWH.

## Acknowledgments

We thank the Program For Advanced Research Capacities For Aids In Peru (Paracas) team and the HIV program personnel from Cayetano Heredia Hospital for their support. Fernando Mejía, MD provided guidance in data collection. Sten Vermund reviewed the manuscript.

## Data availability statement

Due to the sensitive nature of some of the data, including that related to children, data included in this manuscript have not been placed in an open-access database. However, the data that support the findings of this study are available from the corresponding author upon reasonable request. Inquiries should be directed to the corresponding author.

## Notes

### Competing Interest Statement

The authors have declared no competing interest.

### Funding Statement

Yes

### Author Declarations

Ethics Committee of Universidad Peruana Cayetano Heredia and of the study center

